# Evaluating the Effectiveness of a Digital mHealth Intervention (Bei App) to Support Multimodal Treatment in Patients with Fibromyalgia: A Randomized Controlled Trial Protocol

**DOI:** 10.1101/2025.07.29.25330782

**Authors:** Di-Bonaventura Silvia, Ferrer-Peña Raúl, Gurdiel-Álvarez Francisco, Mercado Francisco, Fierro-Marrero José, Morales-Tejera David, Riquelme-Aguado Víctor, Pacheco-Barrios Kevin

**Affiliations:** Department of Physical Therapy, Occupational Therapy, Rehabilitation and Physical Medicine, Rey Juan Carlos University, Alcorcon, Spain; Cognitive Neuroscience, Pain and Rehabilitation Research Group (NECODOR), Faculty of Health Sciences, Rey Juan Carlos University, Madrid, Spain; Clinical and Teaching Research Group on Rehabilitation Sciences (INDOCLIN), CSEU La Salle, UAM, 28023 Madrid, Spain; Centro Superior de Estudios Universitarios La Salle (CSEU La Salle), Autonomous University of Madrid (UAM), 28049 Madrid, Spain; Physiotherapy Unit. Entrevías Health Centre. Primary Care Assistance Management. Madrid Health Service. Madrid. Madrid. Spain; Department of Psychology, Faculty of Health Sciences, Universidad Rey Juan Carlos, Av. Atenas s/n, Madrid 28922, Spain; FENNSI Group, Hospital Nacional de Parapléjicos, SESCAM, Finca la Peraleda, s/n, Toledo 45004, Spain; Motion in Brains Research Group, Centro Superior de Estudios Universitarios La Salle, 28023 Madrid (Spain); Department of Physiotherapy, Human Physiology and Anatomy, Faculty of Physical Education and Physiotherapy, Vrije Universiteit Brussel, Laarbeeklaan 103, 1090 Brussels, Belgium; Pain in Motion Research Group (PAIN), Department of Physiotherapy, Human Physiology and Anatomy, Faculty of Physical Education and Physiotherapy, Vrije Universiteit Brussel, 1090 Brussels, Belgium; Department of Basic Health Sciences, Rey Juan Carlos University, 28933 Madrid, Spain; Grupo de Investigación Consolidado de Bases Anatómicas, Moleculares y del Desarrollo Humano de la Universidad Rey Juan Carlos (GAMDES), 28922 Alcorcón, Spain; Neuromodulation Center and Center for Clinical Research Learning, Spaulding Rehabilitation Hospital and Massachusetts General Hospital, Harvard Medical School; Vicerrectorado de Investigación, Unidad de Investigación para la Generación y Síntesis de Evidencias en Salud, Universidad San Ignacio de Loyola

## Abstract

**Background:** Fibromyalgia (FM) is a complex, chronic condition characterized by widespread musculoskeletal pain, cognitive dysfunction, fatigue, and emotional comorbidities such as anxiety and depression. These symptoms severely impact daily functioning and quality of life. Although non-pharmacological strategies—such as therapeutic exercise and patient education—are recommended as first-line treatments, their implementation outside the clinical setting remains a challenge due to low adherence and lack of continuity.

**Objective:** To evaluate the effectiveness of a digital health intervention—BEI app—as a complement to usual care in improving clinical and functional outcomes among patients with fibromyalgia.

**Methods:** This is a randomized controlled trial (RCT) with two parallel arms (1:1 allocation). A total of 70 adult participants with fibromyalgia (diagnosed per ACR 2016 criteria) will be recruited through a patient association in Madrid, Spain. The control group will receive 12 weeks of standard, in-person treatment consisting of group-based education and physical activity. The experimental group will receive the same in-person program plus daily access to the BEI mobile application, which includes educational modules, physical and cognitive training, symptom tracking, and personalized feedback. Outcomes will be assessed at baseline, week 6 (mid-intervention), week 12 (post-intervention), and week 24 (follow-up). Primary outcomes include pain intensity (VAS) and functional impact (FIQR). Secondary outcomes include catastrophizing, self-efficacy, cognitive function, physical activity, anxiety, depression, quality of life, app engagement, and satisfaction.

**Expected results:** The results of this trial will provide evidence regarding the potential of mHealth tools to enhance adherence, self-efficacy, and functional outcomes in individuals with fibromyalgia.

**Trial registration:** NCT07090434 (ClinicalTrials.gov)

## Introduction

Fibromyalgia (FM) is a prevalent chronic pain disorder that poses a significant public health challenge due to its multifactorial symptomatology and high rates of disability (1). Epidemiological data suggest that FM affects between 2–4% of the general population and up to 6% in primary care, predominantly affecting women (2,3). The constellation of symptoms—chronic pain, severe fatigue, poor sleep, cognitive dysfunction, anxiety, and depression—results in a profound reduction in patients’ quality of life and occupational participation (4).

Clinical guidelines from organizations such as the European League Against Rheumatism (EULAR) emphasize a multimodal and non-pharmacological approach to treatment (5). This includes patient education, structured physical exercise, and cognitive behavioral interventions, which are considered foundational. Pharmacological treatment is only recommended as a supportive strategy due to modest efficacy and common side effects (6).

Despite these recommendations, translating this model into practice is difficult. Public health systems often lack the resources to offer long-term, interdisciplinary care (7). Patients frequently report difficulty in maintaining self-care routines outside of supervised environments, and adherence to lifestyle changes often declines after treatment ends (8). As a result, many patients are left to manage their condition without adequate support. Digital health interventions, especially mobile health (mHealth) applications, offer promising solutions to these gaps (9). Apps can extend care between clinical visits by providing structured education, exercise programs, daily tracking of symptoms, and automated motivational feedback. However, many existing apps are not clinically validated, are not integrated with existing care pathways, and often adopt a generic, one-size-fits-all approach that does not consider the heterogeneity of FM symptoms or user profiles (10).

To address these limitations, Bei app was developed through a co-design process involving patients, clinicians, and digital health experts. It includes tailored educational modules, physical and cognitive exercises, personalized daily tracking symptoms, and goal-setting tools. The goal is to improve long-term adherence, empower patients in their self-care, and complement existing treatments. This protocol describes a randomized controlled trial designed to evaluate the clinical effectiveness of Bei app when used alongside an in-person psychoeducational program for fibromyalgia patients.

## Objectives

### Primary Objectives

1. To determine whether the addition of Bei app to a structured in-person treatment program of psychoeducation results in greater reduction in pain intensity, as measured by the Visual Analogue Scale (VAS), compared to treatment alone.
2. To evaluate whether this intervention leads to significant improvement in functional impact, assessed using the Revised Fibromyalgia Impact Questionnaire (FIQR).

### Secondary Objectives

1. To assess whether the intervention leads to improvement in psychological variables such as pain catastrophizing (PCS), self-efficacy (PSEQ), anxiety and depression (HADS), and kinesiophobia (TSK-11).
2. To evaluate cognitive function improvements using the MoCA and changes in self-reported physical activity via IPAQ-SF.
3. To monitor the daily evolution of symptoms (e.g., pain, fatigue, emotional state) using in-app logging features.
4. To examine the level of adherence to the app (frequency, duration, completion of logs and educational modules).
5. To assess the knowledge acquisition via in-app quizzes and the satisfaction with the digital intervention.

## Methods

### Study Design

This study is designed as a two-arm, randomized, parallel-group, controlled clinical trial. Participants will be randomly allocated (1:1) to either a control group receiving conventional treatment (psychoeducation program) or an experimental group receiving the same treatment plus access to Bei app. The intervention will last 12 weeks, followed by a 12-week observational follow-up, totaling 24 weeks per participant.

The trial adheres to the guidelines of the SPIRIT 2013 and CONSORT statements for protocol reporting and RCT transparency. It is prospectively registered at ClinicalTrials.gov (NCT07090434) and has been approved by the Ethics Committee of Rey Juan Carlos University (URJC) with internal reference number 110620255252025.

### Study Setting and Participant Eligibility

#### Study Setting

Participants will be recruited through the AFISYNFACRO association in Móstoles (Madrid, Spain), which offers structured psychoeducation and exercise programs for people with fibromyalgia.

#### Inclusion Criteria

Participants will be adults between 18 and 65 years of age with a clinical diagnosis of fibromyalgia according to the 2016 criteria established by the American College of Rheumatology (ACR). All participants must be actively enrolled in the group sessions offered by the patient association and possess the ability to use a smartphone or tablet compatible with the BEI app. Informed consent must be provided voluntarily. Additionally, participants are required to have a minimum level of digital health literacy, operationalized as a score equal to or greater than 2.5 in at least four dimensions of the eHealth Literacy Questionnaire (eHLQ).

#### Exclusion Criteria

Participants will be excluded if they present with severe or unstable psychiatric disorders, such as schizophrenia or uncontrolled major depression. Other exclusion criteria include medical comorbidities that contraindicate physical activity, current pregnancy, or concurrent participation in another digital health program or the use of similar therapeutic applications. Individuals with cognitive impairments or technological limitations that prevent the use of the app or participation in digital assessments will also be excluded.

### Recruitment and Consent Process

Recruitment will be conducted via posters, direct communication from association staff, and informative sessions held at AFISYNFACRO facilities. Interested individuals will be pre-screened and complete the eHLQ to assess digital literacy. Eligible participants will be invited to an individual baseline session where the study protocol will be explained in detail and informed consent will be obtained.

### Interventions

#### Control Group (Usual Care)

Participants will attend **12 weekly group sessions**, each lasting one hour, focused on psychoeducation. Sessions are delivered in person by trained professionals (physiotherapists and psychologists) from the association.

#### Experimental Group (Usual Care + Bei App)

In addition to the above, participants will receive access to the **BEI mobile app**, designed to reinforce education and exercise strategies outside clinical sessions. Participants will be instructed to use the app **daily**, guided by a to-do list including:

- 1 educational module (interactive format) + 3 questions self-exam
- 1 physical activity (video-guided, adapted by fatigue and pain level)
- 1 gamified cognitive task (for memory, attention, speed processing and/or executive function)
- Daily mood and symptom check-in

Bei integrates adaptive feedback based on usage patterns, providing a more personalized experience to support long-term adherence.

## Outcomes and Measurement Instruments

### Primary Outcomes

#### Pain Intensity

Pain intensity will be measured using a 100 mm Numeric Rating Scale (NRS), where 0 represents “no pain” and 100 represents the “worst pain imaginable.” Participants will mark a point on the line that best reflected the pain they were experiencing at the time of measurement. Higher scores indicated higher levels of pain, and the administration required less than one minute (11).

#### Functional Impact

It will be measured using the Revised Fibromyalgia Impact Questionnaire (FIQR), a tool designed to assess the impact of fibromyalgia on patients’ daily lives. The FIQR consists of 21 items that evaluate various dimensions, including physical functioning, symptom severity, general health, and pain perception. Items are scored on a scale from 0 to 10, with higher scores indicating a greater impact of the disease. It has demonstrated high internal consistency, with a Cronbach’s alpha ranging from 0.91 to 0.95 (12).

### Secondary Outcomes

#### Pain Catastrophizing

It will be assessed using the Pain Catastrophizing Scale (PCS), which measures the tendency to think catastrophically about pain. The scale consists of 13 items covering three dimensions: rumination, magnification, and helplessness in the face of pain. A Likert scale from 0 to 4 is used, with higher scores indicating greater levels of catastrophizing. The PCS has demonstrated test-retest reliability of 0.84 and adequate internal consistency, with a Cronbach’s alpha of 0.79 (13).

#### Self-Efficacy

It will be assessed using the Pain Self-Efficacy Questionnaire (PSEQ), a self-report questionnaire consisting of 10 items, designed to measure an individual’s self-efficacy in managing pain. It assesses the patient’s confidence in their ability to perform daily activities despite pain, reflecting their sense of control and autonomy in the face of painful situations. The PSEQ has demonstrated excellent internal consistency, with a Cronbach’s alpha of 0.92, and high test-retest reliability with an ICC of 0.90 (14).

#### Cognitive Function

It will be measured using the Montreal Cognitive Assessment (MoCA), a screening tool designed to quickly assess cognitive function in adults. The MoCA consists of 30 items that evaluate various cognitive domains, including attention, memory, language, visuospatial abilities, and executive functions. The test takes approximately 10–15 minutes to complete and is particularly useful in both clinical and research settings for a rapid and effective evaluation of cognitive functioning. The MoCA has demonstrated good validity and reliability across various populations, with a Cronbach’s alpha of 0.77 and an intra-rater correlation of 0.92 (15).

#### Physical Activity

It will be assessed using the International Physical Activity Questionnaire (IPAQ – short version), which is designed to quickly and efficiently measure physical activity levels in adults. It includes questions that evaluate the frequency and intensity of physical activities performed over the past week, including walking, moderate activity, and vigorous activity (16).

#### Kinesiophobia

It will be assessed using a self-report The Tampa Scale for Kinesiophobia (TSK-11), a self-report questionnaire comprising 11 items. The internal consistency of the TSK is high, with Cronbach’s alpha coefficients ranging from 0.74 to 0.93, indicating strong reliability. Test-retest reliability is also good, with correlation coefficients ranging from 0.75 to 0.88 (17).

#### Anxiety and Depression

It will be assessed using the validated Spanish version of the Hospital Anxiety and Depression Scale (HADS), which is divided into two subscales of 7 items each: 1) Depression (HADS-Dep); and 2) Anxiety (HADS-Anx). The subscales of HADS showed internal consistency indices recommended for screening tools. The items in HADS demonstrated a positive correlation with the total score of the anxiety and depression subscales. HADS was found to perform well in assessing the symptom severity and caseness of anxiety disorders and depression in both somatic, psychiatric, and primary care patients and in the general population (18).

#### Quality of Life

It will be measured with the EuroQoL-5D (EQ-5D) questionnaire, a self-report instrument for assessing health-related quality of life. It comprises three elements: a descriptive scale of 5 factors, a second element composed of a vertical NRS, and a social value index generated by the instrument. The EQ-5D has shown good psychometric properties (19).

#### Digital Literacy (baseline only)

Using 4 dimensions of the **eHLQ** as a screening tool (20).

#### App Engagement

Frequency of app use, session length, completion of logs and modules (automatically recorded).

#### Knowledge and Satisfaction

In-app quizzes and a satisfaction questionnaire at week 12.

## Assignment of Interventions

### Randomization Procedure

Participants will be randomly assigned (1:1) to either study arm using a computer-generated randomization list with variable block sizes of 4. The list will be created by an independent statistician to ensure allocation concealment and minimize selection bias.

Allocation concealment will be ensured using sequentially numbered, opaque, sealed envelopes prepared by a researcher not involved in participant interaction. Envelopes will only be opened once the baseline assessment is complete.

### Blinding

Due to the nature of the intervention, which involves the use of a digital app, it will not be possible to blind participants or therapists. However, several procedures will be implemented to reduce potential bias. Outcome assessors will remain blinded to group allocation throughout the study. Statistical analyses will be conducted using pseudonymized datasets, with participants identified only by unique ID codes. Additionally, researchers involved in the development of the app will not take part in outcome assessments or data analysis, ensuring the independence of these processes.

## Statistical Methods

### Descriptive and Comparative Analysis

Data analysis will be conducted using the statistical software SPSS version 25.0 (SPSS Inc., Chicago, IL, USA). A descriptive analysis of the demographic characteristics and pain intensity of the sample size will be performed, presenting continuous variables as mean ± standard deviation (SD) and 95% confidence interval (CI), while categorical variables will be presented as number (n) and percentage (relative frequency, %). In instances where quantitative variables follow a non-normal distribution, they will be described using the median and interquartile range. For the comparison between groups, parametric tests based on the central limit theorem will be chosen, since the sample size of both groups will be greater than 30 (21). A mixed model Analysis of Covariance (ANCOVA) will be used including baseline as the covariate, to obtain between-group adjusted mean differences at each follow-up point, controlling for type I error rate using Bonferroni’s correction. The chi-square test will be used for nominal variables. If the criteria for parametric testing cannot be met, robust methods will be applied (22). A pvalue < 0.05 will be accepted as statistically significant.

### Handling Missing Data

Missing values will be handled using maximum likelihood estimation (MLE). A sensitivity analysis will be conducted using multiple imputation if >10% of any primary outcome is missing.

### Effect Sizes and Significance

Statistical significance will be set at an alpha level of 0.05 (two-tailed). Effect sizes will be calculated using Cohen’s *d* to quantify the magnitude of between-group differences. These will be interpreted according to conventional thresholds, with values of 0.2 considered small, 0.5 medium, and 0.8 or greater indicating a large effect.

### Software

Data will be analyzed using R (version 4.4.0).

## Data Availability

All data produced in the present study are available upon reasonable request to the authors

## Ethics and Dissemination

### Ethical Approval and Registration

This study received ethical approval from the Ethics Committee of Rey Juan Carlos University (Madrid, Spain) (Internal n. registration: 110620255252025). All procedures involving human participants will be conducted in accordance with the ethical standards of the institutional research committee, the principles of the Declaration of Helsinki, and current European data protection regulations (GDPR). The trial has been prospectively registered at ClinicalTrials.gov under the identifier NCT07090434.

### Informed Consent

Participants will receive written and oral explanations of the study. Informed consent will be obtained prior to any study procedures. Consent forms include details on voluntary participation, confidentiality, and data use.

### Data Protection and Confidentiality

Data will be stored in the REDCap system, encrypted and password protected. Each participant will be assigned a code, and identifying information will be kept separate. Data will be retained for five years post-study, then securely deleted per institutional and legal regulations.

### Adverse Events and Monitoring

Due to the low-risk nature of the intervention, no external data monitoring board is required. Any adverse events or protocol deviations will be documented and reviewed internally by the PI and research team.

### Dissemination

Results will be published in **peer-reviewed open-access journals** and presented at scientific conferences. Summaries will be provided to participants upon request. Anonymized datasets and analysis scripts will be available under request for replication or meta-analysis.

## References

1. Sarzi-Puttini P, Giorgi V, Marotto D, Atzeni F. Fibromyalgia: an update on clinical characteristics, aetiopathogenesis and treatment. Nat Rev Rheumatol [Internet]. 2020 Nov 1 [cited 2025 Jul 1];16(11):645–60. Available from: https://pubmed.ncbi.nlm.nih.gov/33024295/

2. Kocyigit BF, Akyol A. Fibromyalgia syndrome: epidemiology, diagnosis and treatment. Reumatologia [Internet]. 2022 [cited 2025 Jul 1];60(6):413. Available from: https://pmc.ncbi.nlm.nih.gov/articles/PMC9847104/

3. Andrews P, Steultjens M, Riskowski J. Chronic widespread pain prevalence in the general population: A systematic review. European Journal of Pain (United Kingdom) [Internet]. 2018 Jan 1 [cited 2025 Jul 1];22(1):5–18. Available from: /doi/pdf/10.1002/ejp.1090

4. Siracusa R, Di Paola R, Cuzzocrea S, Impellizzeri D. Fibromyalgia: Pathogenesis, mechanisms, diagnosis and treatment options update. Int J Mol Sci [Internet]. 2021 Apr 2 [cited 2025 Jul 1];22(8). Available from: https://pubmed.ncbi.nlm.nih.gov/33918736/

5. Macfarlane GJ, Kronisch C, Dean LE, Atzeni F, Häuser W, Flub E, et al. EULAR revised recommendations for the management of fibromyalgia. Ann Rheum Dis [Internet]. 2017 Feb 1 [cited 2025 Jul 1];76(2):318–28. Available from: https://pubmed.ncbi.nlm.nih.gov/27377815/

6. Perrot S. Fibromyalgia: do I tackle you with pharmacological treatments? Pain Rep [Internet]. 2025 Jan 9 [cited 2025 Jul 1];10(1):e1222. Available from: https://journals.lww.com/painrpts/fulltext/2025/02000/fibromyalgiado_i_tackle_you_with_pharmacological.17.aspx

7. Fischer L, Smeets RGM, Rijken M, Elissen AMJ. Barriers and facilitators to integrated primary care from the perspective of people with chronic conditions and multiple care needs: A scoping review. Health Policy (New York) [Internet]. 2025 Feb 23 [cited 2025 Jul 1];105283. Available from: https://www.sciencedirect.com/science/article/pii/S0168851025000399

8. Yung K, Jadhav D, Ma C, Majgaonkar S, Manai E, Pearson J. Exploring patient activation and self-management experiences in adults with fibromyalgia: a qualitative evidence synthesis. Rheumatol Adv Pract [Internet]. 2025 Mar 14 [cited 2025 Jul 1];9(2). Available from: https://dx.doi.org/10.1093/rap/rkaf025

9. Donisi V, De Lucia A, Pasini I, Gandolfi M, Schweiger V, Del Piccolo L, et al. e-Health Interventions Targeting Pain-Related Psychological Variables in Fibromyalgia: A Systematic Review. Healthcare [Internet]. 2023 Jul 1 [cited 2025 Jul 1];11(13):1845. Available from: https://pmc.ncbi.nlm.nih.gov/articles/PMC10341245/

10. An J, Fan W, Mittal A, Zhang Y, Chen AT. Mobile App Use among Persons with Fibromyalgia: A Cross-sectional Survey. J Pain [Internet]. 2024 Aug 1 [cited 2025 Jul 1];25(8):104515. Available from: https://www.sciencedirect.com/science/article/abs/pii/S1526590024004358

11. Hjermstad MJ, Fayers PM, Haugen DF, Caraceni A, Hanks GW, Loge JH, et al. Studies comparing numerical rating scales, verbal rating scales, and visual analogue scales for assessment of pain intensity in adults: A systematic literature review. Vol. 41, Journal of Pain and Symptom Management. 2011. p. 1073–93.

12. Salgueiro M, García-Leiva JM, Ballesteros J, Hidalgo J, Molina R, Calandre EP. Validation of a Spanish version of the Revised Fibromyalgia Impact Questionnaire (FIQR). Health Qual Life Outcomes [Internet]. 2013 Aug 1 [cited 2025 Jul 2];11(1). Available from: https://pubmed.ncbi.nlm.nih.gov/23915386/

13. García Campayo J, Rodero B, Marta Alda M, Sobradiel N, Montero J, Moreno S. Validación de la versión española de la escala de la catastrofización ante el dolor (Pain Catastrophizing Scale) en la fibromialgia. Med clín (Ed impr). 2008;487–93.

14. Perez-Dominguez B, Perpiña-Martinez S, Escobio-Prieto I, de la Fuente-Costa M, Rodriguez-Rodriguez AM, Blanco-Diaz M. Psychometric properties of the translated Spanish version of the Pain Self-Efficacy Questionnaire. Front Med (Lausanne). 2023 Jul 3;10:1226037.

15. Delgado C, Araneda A, Behrens MI. Validación del instrumento Montreal Cognitive Assessment en español en adultos mayores de 60 años. Neurología [Internet]. 2019 Jul 1 [cited 2025 Jul 2];34(6):376–85. Available from: https://repositorio.uchile.cl/handle/2250/172983

16. Craig CL, Marshall AL, Sjöström M, Bauman AE, Booth ML, Ainsworth BE, et al. International physical activity questionnaire: 12-Country reliability and validity. Med Sci Sports Exerc [Internet]. 2003 Aug 1 [cited 2025 Jul 2];35(8):1381–95. Available from: https://pubmed.ncbi.nlm.nih.gov/12900694/

17. Woby SR, Roach NK, Urmston M, Watson PJ. Psychometric properties of the TSK-11: A shortened version of the Tampa Scale for Kinesiophobia. Pain [Internet]. 2005 Sep [cited 2025 Jul 2];117(1–2):137–44. Available from: https://journals.lww.com/pain/fulltext/2005/09000/psychometric_properties_of_the_tsk_11a_shortened.16.aspx

18. Bjelland I, Dahl AA, Haug T, Neckelmann D. The validity of the Hospital Anxiety and Depression Scale An updated literature review.

19. Cabasés JM. The EQ-5D as a measure of health outcomes. Gac Sanit. 2015 Nov 1;29(6):401–3.

20. Kayser L, Karnoe A, Furstrand D, Batterham R, Christensen KB, Elsworth G, et al. A multidimensional tool based on the eHealth Literacy Framework: Development and initial validity testing of the eHealth Literacy Questionnaire (eHLQ). J Med Internet Res [Internet]. 2018 Feb 1 [cited 2025 Jul 2];20(2). Available from: https://pubmed.ncbi.nlm.nih.gov/29434011/

21. Kwak SG, Kim JH. Central limit theorem: the cornerstone of modern statistics. Korean J Anesthesiol [Internet]. 2017 Apr 1 [cited 2025 Jul 2];70(2):144. Available from: https://pmc.ncbi.nlm.nih.gov/articles/PMC5370305/

22. Vrbin CM. Parametric or nonparametric statistical tests: Considerations when choosing the most appropriate option for your data. Cytopathology [Internet]. 2022 Nov 1 [cited 2025 Jul 2];33(6):663–7. Available from: https://pubmed.ncbi.nlm.nih.gov/36017662/

